# Values and Preferences Related to Workplace Mental Health Programs and Interventions: An International Survey

**DOI:** 10.1101/2023.03.02.23286684

**Authors:** J.K. Murphy, J.M. Noble, P.A. Chakraborty, G. Michlig, E.E Michalak, A.J. Greenshaw, R.W. Lam

**Author notes:** (co-first authors).

## Abstract

**Introduction:** This study explores the perspectives of workers and managers on workplace programs and interventions that seek to promote mental wellbeing, and prevent and treat mental health conditions The results contributed supporting evidence for the development of the WHO’s first global guidelines for mental health and work, which provide evidence-based recommendations to support the implementation of workplace mental health programs and supports, to improve their acceptability, appropriateness, and uptake.

**Methods:** A survey was used to examine the values and preferences among workers and employers related to workplace mental health prevention, protection, promotion, and support programs and services. The survey was made available in English, French, and Spanish. Descriptive statistics were used to analyse the survey data. Rapid thematic analysis was used to analyse the results of qualitative questions.

**Results:** These results provide a unique international perspective on programs and supports for mental health at work, from the standpoint of employees, including managers. Results suggest that employees value interventions developed in consultation with workers (including indicated, selective and universal interventions), increased training and capacity building among managers, and targeted interventions to address the pervasive impact of stigma on perceptions about mental health at work and help-seeking. The findings of this study seek to reflect the perspectives of workers, including managers, and therein to promote improved access, availability and uptake of mental health programs and supports at work and – ultimately- to support the potential of workplaces as environments that promote and support mental health.

## Introduction and Background

Mental health conditions, including substance-use disorders, are prevalent worldwide and lead to a substantial burden on individuals and high socioeconomic cost. Globally, mental health conditions are a leading cause of illness and disability (1, 2). Left untreated, they are associated with poor physical health (3), reduced quality of life (4), lower functional capacity (5) and lost productivity (6). People living with mental health conditions may also experience stigma, discrimination and human rights violations (7). Despite the risks associated with poor mental health, approximately 50% of people worldwide do not have access to evidence-based mental health support, and in the lowest resourced countries this gap may be up to 90% (8).

Work and mental health are inextricably linked. Work may have a positive or negative impact on mental health, while mental ill health may affect work performance, productivity, and employment status (9). Mental health conditions affecting workers include clinically diagnosed conditions like depression, anxiety or alcohol use disorders and sub-clinical issues including workplace stress and burnout (10). Experiences of mental ill health may lead to negative impacts for workers including workplace discrimination (10). The impact of untreated mental health conditions for workplaces include the costs of absenteeism (time off from work due to illness), presenteeism (reduced work capacity due to working while ill), lost productivity, increased risk of workplace injury or safety incidents, disability claims, and challenges with retention (11, 12). It is estimated that in the world’s 36 largest countries, the loss of workplace productivity due to untreated depression and anxiety is equivalent to 12 billion work days each year, costing up to US $925 billion (13).

People over the age of 18 years spend more than 60% of waking hours at work (14), suggesting that the workplace presents a considerable opportunity for the provision of mental health prevention, protection, promotion and support interventions. Considerable savings and financial benefit may result from investing in workplace mental health promotion and prevention (15). Despite this, access to occupational health services remains low worldwide, with only 20-50% of workers in high income settings and 5-10% in low and middle-income countries (LMICs) able to access to such supports (16, 17).

Understanding the perspectives and experiences of workers is essential to informing how work and workplaces can best provide appropriate, targeted support to promote workforce mental health and wellbeing. This survey assessed the values and preferences of workers and employers related to workplace programs and interventions to promote mental wellbeing and prevent and treat mental health conditions. This survey was commissioned by the World Health Organization (WHO) to provide supporting evidence for the development of the WHO’s Guidelines on mental health and work (18).

## Methods

### Survey scope and development

This survey examined values and preferences among workers and employers related to workplace mental health promotion, prevention and support programs and services. The survey explored the following themes: (1) Values and preferences related to mental health and work generally, including help-seeking preferences for programs and services, and; (2) awareness, access, and values and preferences related to specific intervention types. Specific intervention types are described in Box 1. The survey consisted primarily of quantitative questions and we included short-answer, qualitative questions as part of the ‘general values and preferences’ section as described below.

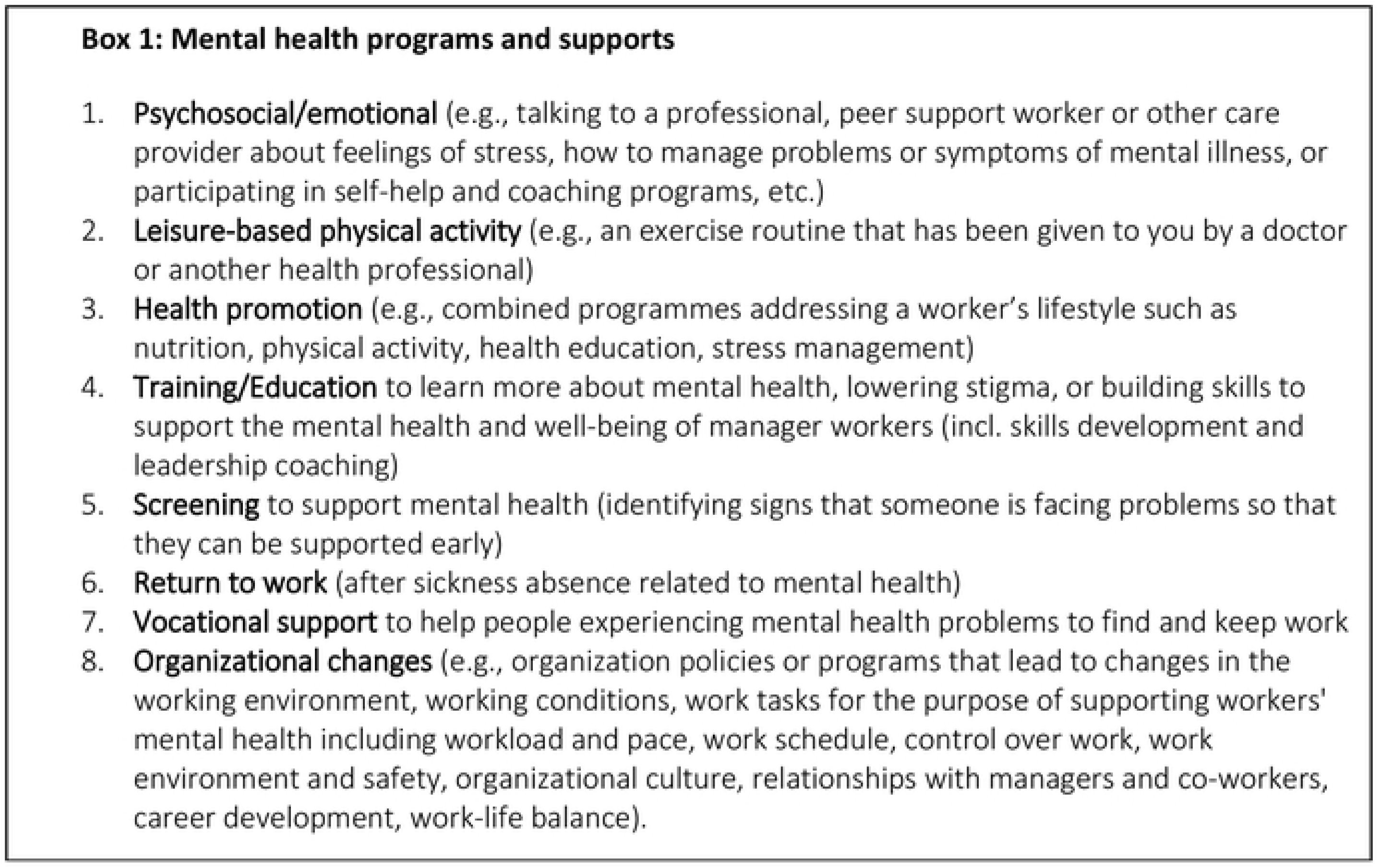

The survey contained specific questions for managers, people who provide mental health support at work, and ‘high-risk’ workers including first responders, humanitarian and healthcare workers. Qualitative questions were included to provide a more comprehensive understanding of attitudes related to mental health program and service provision at work in general, regarding the changing nature of work and mental health, the impact of the COVID-19 pandemic on work and mental health and attitudes related to mental health screening at work. For brevity, this paper focuses on the two themes identified above, with additional data available in the *WHO Web Annex: Evidence profiles and supporting evidence* (Available at: https://apps.who.int/iris/bitstream/handle/10665/363102/9789240053076-eng.pdf).

The survey focused on interventions delivered at or in relation to work that are provided at an individual, manager/ team, and organizational level. Interventions of interest include universal promotion and prevention programs (for the general worker population), selected promotion and prevention programs (for workers in high-risk occupations), indicated promotion and prevention programs (for workers showing signs of sub-threshold emotional distress), and programs to support treatment and recovery (for workers with symptoms of mental health or substance use conditions). The survey also examined perspectives on screening for mental health in the workplace. The full survey is available upon request from the lead authors.

The survey was piloted by ten individuals representing a diversity of backgrounds and from several countries (Canada, the United States, Japan, Zambia, Peru, and the Netherlands), as well as several WHO- affiliated experts and expert groups. We asked pilot participants to provide feedback about their ability to understand the survey questions and terminology, the survey flow, appropriateness of the questions, and survey length. Pilot participant feedback was applied to the final survey, including by removing repetitive questions, providing clear definitions of terminology, and rephrasing some questions for clarity.

Ethics approval for this study was obtained from the University of Alberta’s Research Ethics Office (approval number: Pro00107550). Participants were required to provide written informed consent by by selecting “I consent to participate in this survey” in order to access the survey. Participants were provided with the option to withdraw consent at any time while completing the survey by withdrawing from the survey before submission. All data from withdrawn surveys has been excluded from the final dataset.

### Survey dissemination

This survey was disseminated online in English, French and Spanish using Qualtrics survey software (19). Inclusion criteria for survey participation were employed civilian (non-military) adults over 18 years who are engaged in paid formal, non-standard work or informal work. The survey was open to people worldwide.

The survey was disseminated broadly via several channels, including through international global mental health networks, university networks, via mental health, workplace, and patient-serving organizations, and via social media. Data collection took place between April 9^th^ and May 16^th^, 2021.

### Analysis

We used descriptive statistics to analyse the survey data. Qualitative responses were analysed using rapid thematic analysis (20). Lead author JKM conducted the qualitative analysis, reviewing the responses for immersion in the data and developing a code book based on each question category, with new codes added as they were identified in the dataset. Data were coded using NVivo 12 (21) software, and reviewed to identify overarching themes related to each question category.

## Results

451 surveys were included in the final analysis after cleaning data to remove withdrawn surveys (indicating withdrawal of informed consent) and spam responses. Of these, 224 were complete and 227 were partially completed. We have included data from partially completed surveys except in cases where the survey respondents withdrew their consent. We have indicated in the results tables the total number (n) of responses for each question. In cases where participants could select more than one response (“choose all that apply”), we have indicated the total number of respondents and total number of response selections per item. 29 surveys were completed in Spanish, and 29 were completed in French, with the remaining 393 completed in English. Findings related to the general workforce and managers are reported here. Disaggregated findings related to workers in high-risk occupations will be reported elsewhere.

Quantitative and qualitative results are described together where thematically aligned. For qualitative results, illustrative quotations are provided. Quotations are annotated with participant number based on the order in which they completed the survey, whether they are managers (i.e. supervise staff), and their gender.

### Participant Demographics and Types of Work

451 individuals responded to the survey. Respondents were predominantly highly educated (64.2% with a graduate degree), identified as female (67.2%), averaged around 43 years old, and were living in large or medium sized cities (60.5%). 11.5% identified as sexual and gender minority groups, 18.6% identified as ethnic minority groups in their country of residence and 9.3% identified as Indigenous.

**Table 1:** Participant Characteristics.

| Demographic Variable | Percent |
| --- | --- |
| Gender (n=357) | Percent |
| Woman | 67.2 |
| Man | 31.4 |
| Other | 1.1 |
| Prefer not to say | 0.3 |
| Total | 100 |
| Median Age | 42.8 years |
| Community size (n= 348) | Percent |
| Large city (>1 million inhabitants) | 37.9 |
| Medium city (300,000-1 million inhabitants) | 25.6 |
| Small city (100,000 – 300,000 inhabitants) | 13.5 |
| Large town (20,000-100,000 inhabitants) | 10.3 |
| Medium town (1,000-20,000 inhabitants) | 8.3 |
| Small town, village or hamlet (<1000 inhabitants) | 4.3 |
| Total | 100 |
| Highest Level of Formal Education Completed (n=321) | Percent |
| None | 0.3 |
| Primary school | 0 |
| Secondary/high school (including vocational high school) | 4.0 |
| University Undergraduate Degree (e.g., Bachelor Degree) | 28.7 |
| Graduate Degree (e.g., Master or Doctor Degree) | 64.2 |
| Other | 2.8 |
| Total | 100 |
| Identify as Member of Sexual or Gender Minority Group (n=338) | Percent |
| Yes | 11.5 |
| No | 85.5 |
| Prefer not to say | 2.4 |
| Other | 0.6 |
| Total | 100% |
| Identify as Member of Racial or Ethnic Minority (n=323) | Percent |
| Yes | 18.6 |
| No | 78.6 |
| Prefer not to say | 1.5 |
| Other | 1.2 |
| Total | 100 |
| Identify as Member of an Indigenous Group (n=54) | Percent |
| Yes | 9.3 |
| No | 87.0 |
| Prefer not to say | 1.9 |
| Unsure | 1.9 |
| Total | 100 |
| Caregiving Responsibility at Home (n=338) | Percent |
| Yes | 41.7 |
| No | 58.3 |
| Total | 100 |
| Identify as Living with a Disability (n=314) | Percent |
| Yes | 12.7 |
| No | 87.3 |
| Total | 100 |

Just over half (55.3%) of responses came from people residing in high-income countries, 21.4% from upper-middle income, 14.2% from lower-middle income and 8.9% from low-income countries. Survey responses came from all WHO regions, with a majority of responses from the European region (42.4%) and the region of the Americas (28.5%). Additional responses came from the Western Pacific (13.0%), African region (8.9%), the Eastern Mediterranean (4.8%), and South East Asia (2.4%).

Table 2 displays the employment characteristics of respondents, with the majority (58.3%) engaged in paid full-time formal employment.

**Table 2:**
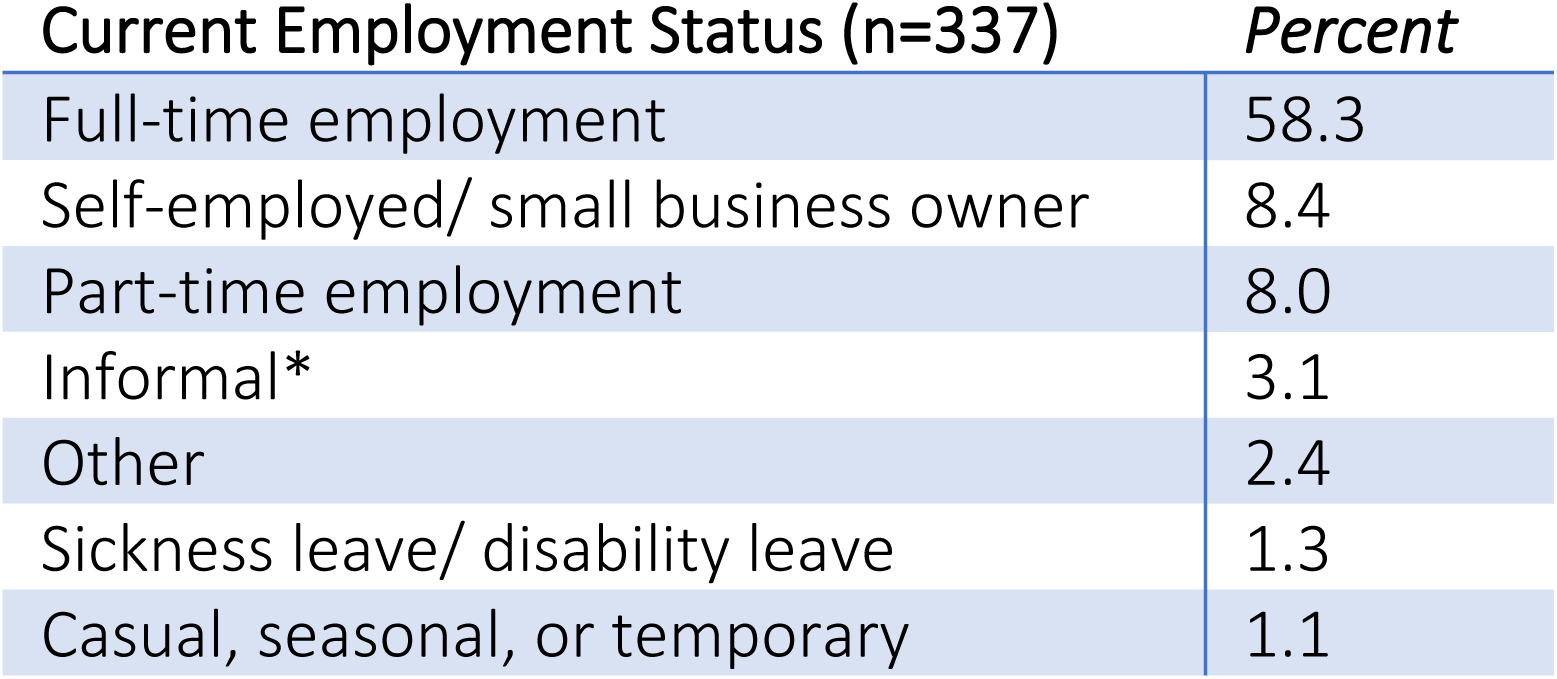
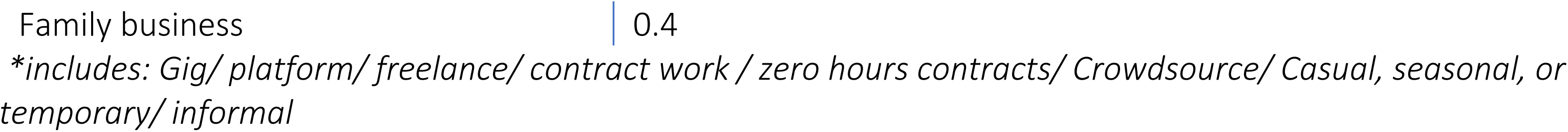
Current Employment Characteristics.

Respondents selected the type of employment in which they are engaged by industry from a drop-down menu based on the United Nations’ International Standard Industrial Classification of All Economic Activities (ISIC) (22). They were able to further select more detail on their employment type via additional drop-down menus and by entering open field data. The majority of respondents are employed in Human Health and Social Work Activities (25.5%), Professional, Scientific and Technical Activities (21.5%) and Education (19.8%). More detail can be found in WHO Web Annex: Evidence profiles and supporting evidence, page. 307 (Available at: https://apps.who.int/iris/bitstream/handle/10665/363102/9789240053076-eng.pdf).

Over half of respondents indicated that they play a role in supporting mental health at work (53.3%) in roles including occupational health and safety, human resources and mental health workers. Just over half of respondents identified themselves as managers (51.9%), indicating that they play a supervisory role at work. A majority of respondents (68.1%) indicate that they have current or prior experience with a mental health condition, and 28.1% have taken a formal leave due to mental health challenges.

### Theme 1: General Values and Preferences Regarding Mental Health at Work

This theme sought to explore respondent’s general beliefs about mental health, and values and preferences regarding mental health programs and supports at work and help-seeking in general.

#### Current Impact

To the question, “What type of impact does your work have generally on your mental health?” respondents reported equally that work has both a positive and negative impact (24.5%) or somewhat positive (24.5%) impact on mental health. Remaining respondents reported that work had a somewhat negative impact on mental health (19.0%), a very positive impact on mental health (15.7%), or a very negative impact (6.2%). 9.8% responded neutrally, and 0.7% were unsure.

We also asked respondents to rate on a six point Likert scale the extent to which several factors, based on established categories of psychosocial risk (23), negatively impact their mental health. As described in Table 3, issues related to “workload and work pace,” including high workload, challenging deadlines and understaffed work environments, had the most negative impact on mental health.

**Table 3:** Psychosocial risk factors perceived negative impact on mental health.

|  | (N=291) | A great deal, % | A lot, % | A moderate amount, % | A little, % | Not at all, % |
| --- | --- | --- | --- | --- | --- | --- |
| Job stability (n=254) |  | 8.3 | 11.8 | 13.4 | 22.1 | 41.3 |
| Work life balance (n=253) |  | 11.1 | 16.2 | 18.2 | 24.5 | 28.1 |
| Career development (n=253) |  | 15.8 | 19.0 | 13.4 | 19.0 | 29.3 |
| Role in organization (n=256) |  | 14.8 | 9.8 | 14.1 | 31.6 | 27.0 |
| Interpersonal relationships at work (n=256) |  | 14.1 | 9.0 | 15.2 | 27.3 | 32.4 |
| Organizational culture and function (n=256) |  | 20.3 | 10.6 | 18.0 | 25.0 | 24.2 |
| Environment and equipment (n=255) |  | 8.6 | 9.4 | 10.6 | 16.9 | 46.7 |
| Job control (n=256) |  | 12.9 | 14.8 | 18.0 | 26.6 | 24.6 |
| Work schedule (n=259) |  | 12.0 | 16.2 | 14.3 | 22.4 | 31.7 |
| Workload and work pace (n=264) |  | 20.8 | 19.3 | 27.7 | 17.8 | 13.3 |
| Work content/ task-design (n=286) |  | 11.5 | 11.2 | 18.2 | 19.6 | 36.7 |

#### Help-Seeking Preferences

We assessed factors related to help-seeking preferences and accessibility of programs and supports among workers. Table 4 describes the comfort level of respondents when speaking about their mental health concerns at work with various individuals, workplace representatives and services.

**Table 4:**
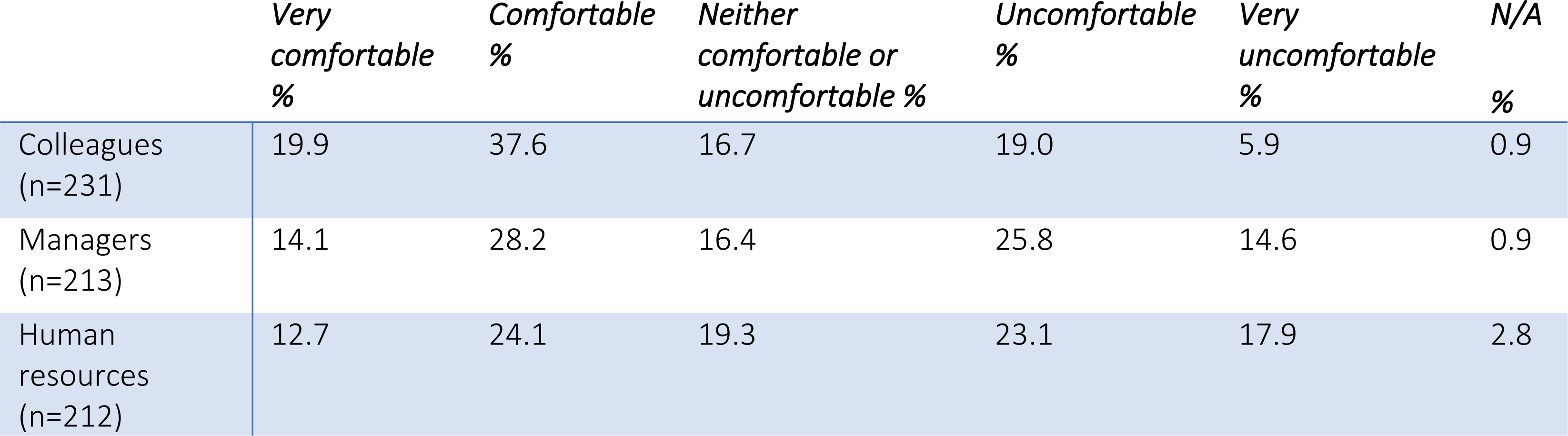
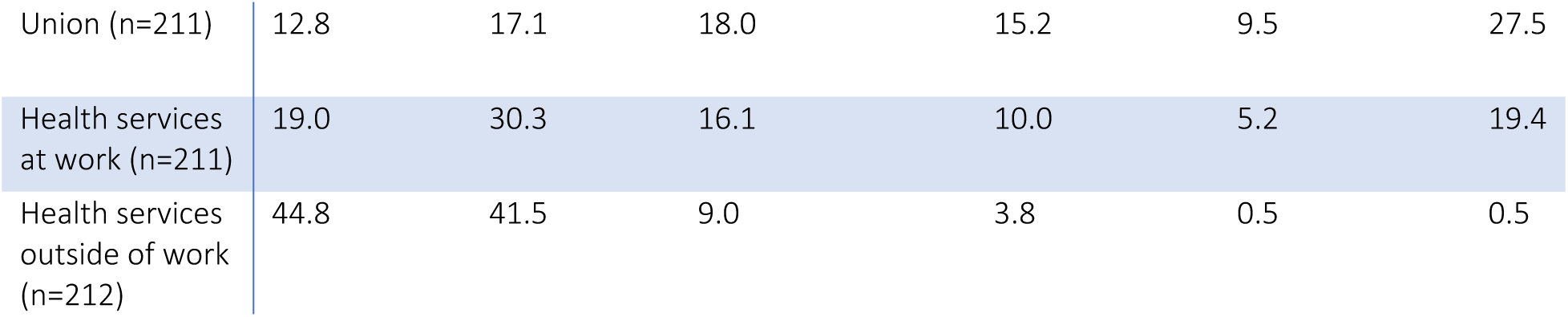
Preferences for speaking about mental health concerns at work.

|  | Very comfortable % | Comfortable % | Neither comfortable or uncomfortable % | Uncomfortable % | Very uncomfortable % | N/A % |
| --- | --- | --- | --- | --- | --- | --- |
| Colleagues (n=231) | 19.9 | 37.6 | 16.7 | 19.0 | 5.9 | 0.9 |
| Managers (n=213) | 14.1 | 28.2 | 16.4 | 25.8 | 14.6 | 0.9 |
| Human resources (n=212) | 12.7 | 24.1 | 19.3 | 23.1 | 17.9 | 2.8 |
| Union (n=211) | 12.8 | 17.1 | 18.0 | 15.2 | 9.5 | 27.5 |
| Health services at work (n=211) | 19.0 | 30.3 | 16.1 | 10.0 | 5.2 | 19.4 |
| Health services outside of work (n=212) | 44.8 | 41.5 | 9.0 | 3.8 | 0.5 | 0.5 |

We sought responses regarding whether respondents had considered participating in mental health programs and supports at work but did not do so and their reasons for not participating so as to elicit potential barriers to participation. 62.6% of respondents had considered participating in an intervention but chose not to. The majority (45.6%) report that they chose not to participate in psychosocial and emotional programs and supports. The reasons for not participating are reported in Table 5.

**Table 5:** Reasons for not participating in mental health programs or supports at work.

| Reasons for Not Participating (n=79; Total responses n=222) | Percent |
| --- | --- |
| Did not have time (n=39) | 17.5 |
| Was not able to access the services or program (n=21) | 9.5 |
| Was worried about privacy (n=25) | 11.3 |
| Preferred to manage by myself(n=30) | 13.5 |
| Did not think it would help/be effective (n=17) | 7.7 |
| Afraid to ask for help from my manager or employer (n=17) | 7.7 |
| Afraid my co-workers would find out (n=10) | 4.5 |
| Did not feel I needed to participate (n=9) | 4.0 |
| Could not afford it (n=5) | 2.2 |
| Lack of choice in service types (n=13) | 5.9 |
| Work is not responsible for supporting or promoting the mental health and wellbeing or workers (n=2) | 0.9 |
| I am too focused on meeting my basic needs (food, clothing, shelter) (n=6) | 2.7 |
| I was worried that asking for help would lead to negative consequences (n=17) | 7.7 |
| Other, please specify* | 4.9 |

\*Other responses: Fear that management would respond negatively to need to take necessary time; feel that most representatives don't have necessary experience and belief that systems is not responsive to needs; Lack of trust in organizers of programs; Work would not provide referral to occupational health; Concerned that the service was not open for specific category of employment

As shown in Table 6, respondents also selected who they would prefer to receive support from across four scenarios related to experiencing mental health challenges at work. Respondents were able to select all that apply from a list of possible resources as shown in Table 6.

**Table 6:** Preferences for seeking support for mental health concerns at work.

| Who would you want support from when... | Colleagues | Managers/<br>supervisors | Human<br>resources | Union<br>reps | Health<br>services at<br>work | Health<br>services<br>outside of<br>work |
| --- | --- | --- | --- | --- | --- | --- |
| First experiencing mental health concerns at work?<br>(n=218/choice count n=451) | 22.4 | 26.4 | 8.0 | 4.0 | 15.7 | 23.5 |
| Seeking ongoing support for mental health at work?<br>(n=49/Choice count n=92) | 14.1 | 25.0 | 10.9 | 6.5 | 20.7 | 22.8 |
| Deciding whether or not to take a leave of absence due to mental health? (n=46/ Choice count n=101) | 5.9 | 28.7 | 18.8 | 5.0 | 16.8 | 24.8 |
| Returning to work after a leave for mental health reasons?<br>(n=44/ Choice count n=113) | 10.6 | 30.1 | 21.2 | 7.1 | 14.2 | 16.8 |

When asked about the importance of accessing mental health supports on their own, without the assistance of involvement of a manager, a large majority (63.5%) of respondents indicated that doing so is very important, not wanting their employer to know they are accessing services or supports for mental health. Responses are displayed in Table 7.

**Table 7:**
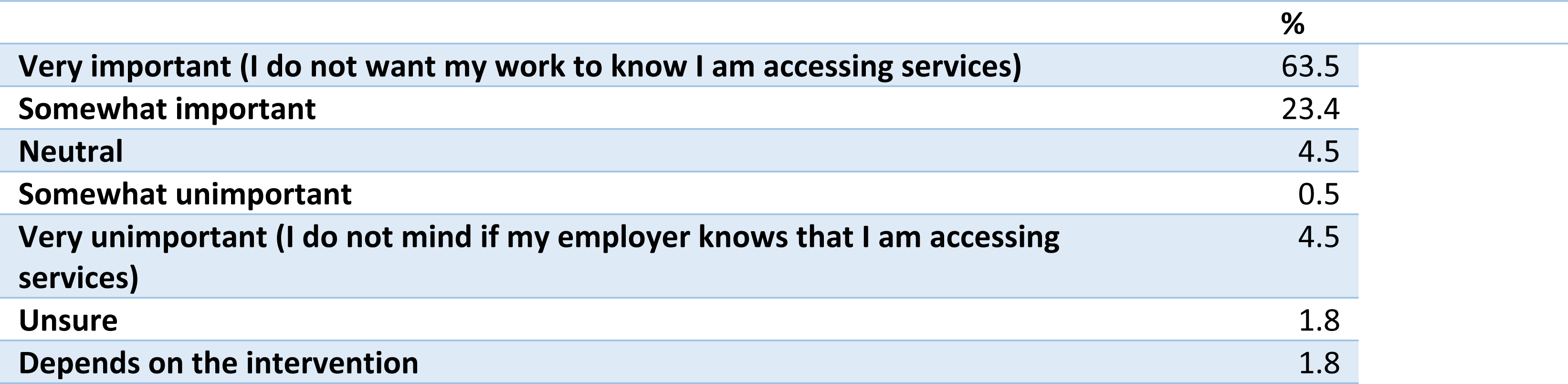
Importance of accessing mental health supports without assistance of manager (*n=222*)

Respondents rated their preferences for how they prefer to access mental health programs if needed (including individual, group, in-person and digital options), where they prefer to access programs or services via the workplace and their preferred method of delivery or provider for mental health support, as shown in Table 8.

**Table 8:** Preferences for mode, location and provider of intervention delivery.

| **Mode (n=210)** | **%** |
| --- | --- |
| **Other<sup>a</sup>** | 1.5 |
| **On my own, digitally (online/ self-directed)** | 32.6 |
| **On my own, in person** | 50.3 |
| **In a group, digitally (online)** | 5.8 |
| **In a group, in person** | 9.8 |
| **Location (n=219)** | **%** |
| **I would not be interested in support** | 0.5 |
| **Other<sup>b</sup>** | 0.5 |
| **Outside of work (not home)** | 42.1 |
| **Online** | 27.7 |
| **At home** | 16.9 |
| **At work** | 12.3 |
| **Provider (n=218)** | **%** |
| **I would not be interested in support** | 0.0 |
| **Other<sup>c</sup>** | 2.7 |
| **No one/ on your own** | 4.0 |
| **Online app or web based program** | 12.9 |
| **Health worker** | 53.0 |
| **Human resources/ occupational health manager** | 14.8 |
| **Manager** | 12.6 |

<sup>a</sup> Not comfortable accessing mental health support at work; on my own over the phone
<sup>b</sup> Home is usually a safe space but during the COVID-19 pandemic this was not the case; the level of psychological safety of location is a priority; a neutral venue (not at work, the home, or an office due to concerns of abuse)
<sup>c</sup> Community-based support; telephone-based; vocational rehab consultant; family, friends, other peer counselors who don't work for my agency; counsellor

We asked respondents to rank from most to least important considerations for employers when offering mental health supports or programs outside of the workplace. Table 9 displays the frequency with which each item was ranked as a top priority.

**Table 9:**
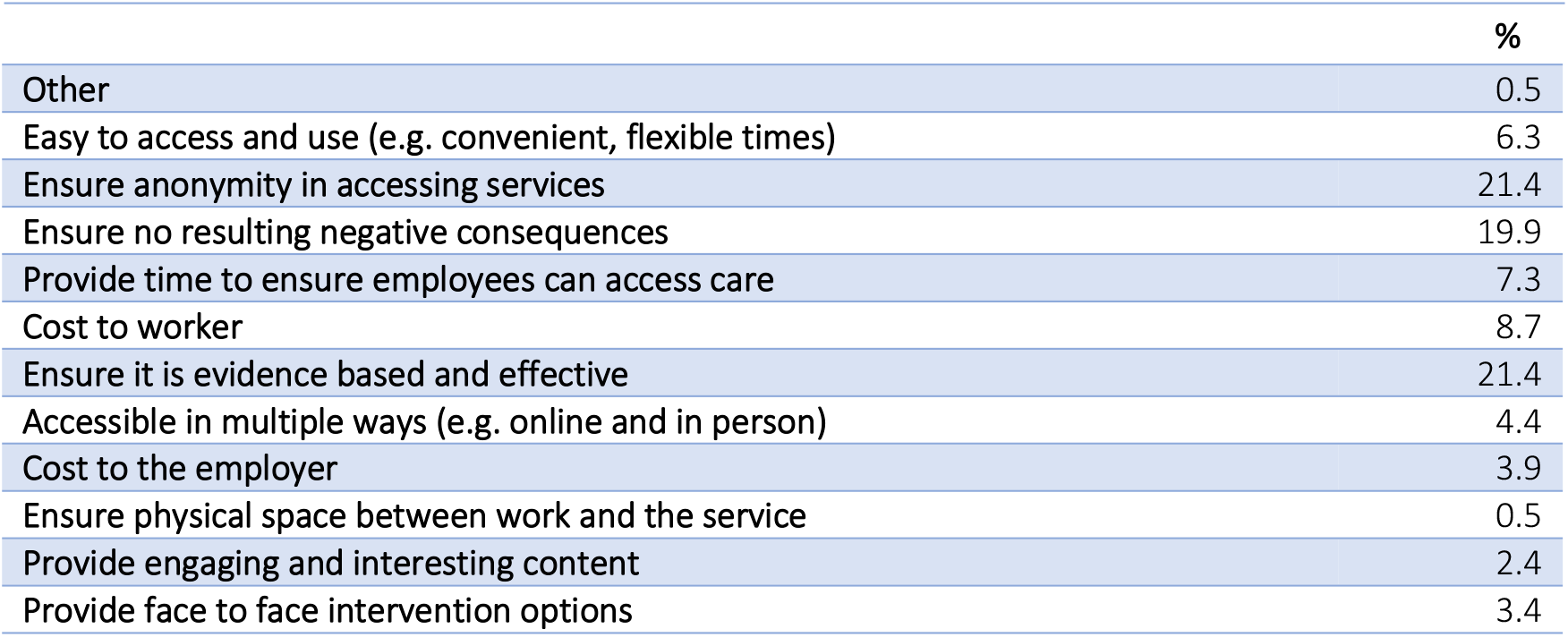
What is most important for employers to consider in offering a mental health service or program that is available to workers outside of work? (n=206)

### Theme 2: Awareness of, access to, and values and preferences for specific intervention types

In this section, the survey sought to explore respondents’ awareness of and access to specific types of mental health programs and supports at work, as well as their values and preferences for those interventions for which they have familiarity. Respondents were asked to provide their perspective on the intervention types described in Figure 1.

#### Awareness of Services and Program Types

We asked respondents to select, from a list of intervention types, all program and service types that they had previously heard of. As shown in Table 10, awareness of service and program types ranges from approximately one half to approximately one third of respondents.

**Table 10:** Respondent awareness of intervention options for mental health at work.

**Interventions (N=248)**
|  |  |
| --- | --- |
| **Psychosocial interventions** | 51.0 |
| **Leisure-based physical activity** | 45.0 |
| **Health promotion** | 46.3 |
| **Training** | 44.1 |
| **Screening programmes** | 32.6 |
| **Return to work** | 38.8 |
| **Vocational support** | 29.3 |
| **Organizational interventions (N=251)** | 60.6 |

Table 11 describes whether respondents have access to each intervention type at work, whether they have previously used each intervention type, and, if not, whether they would be willing to.

**Table 11:**
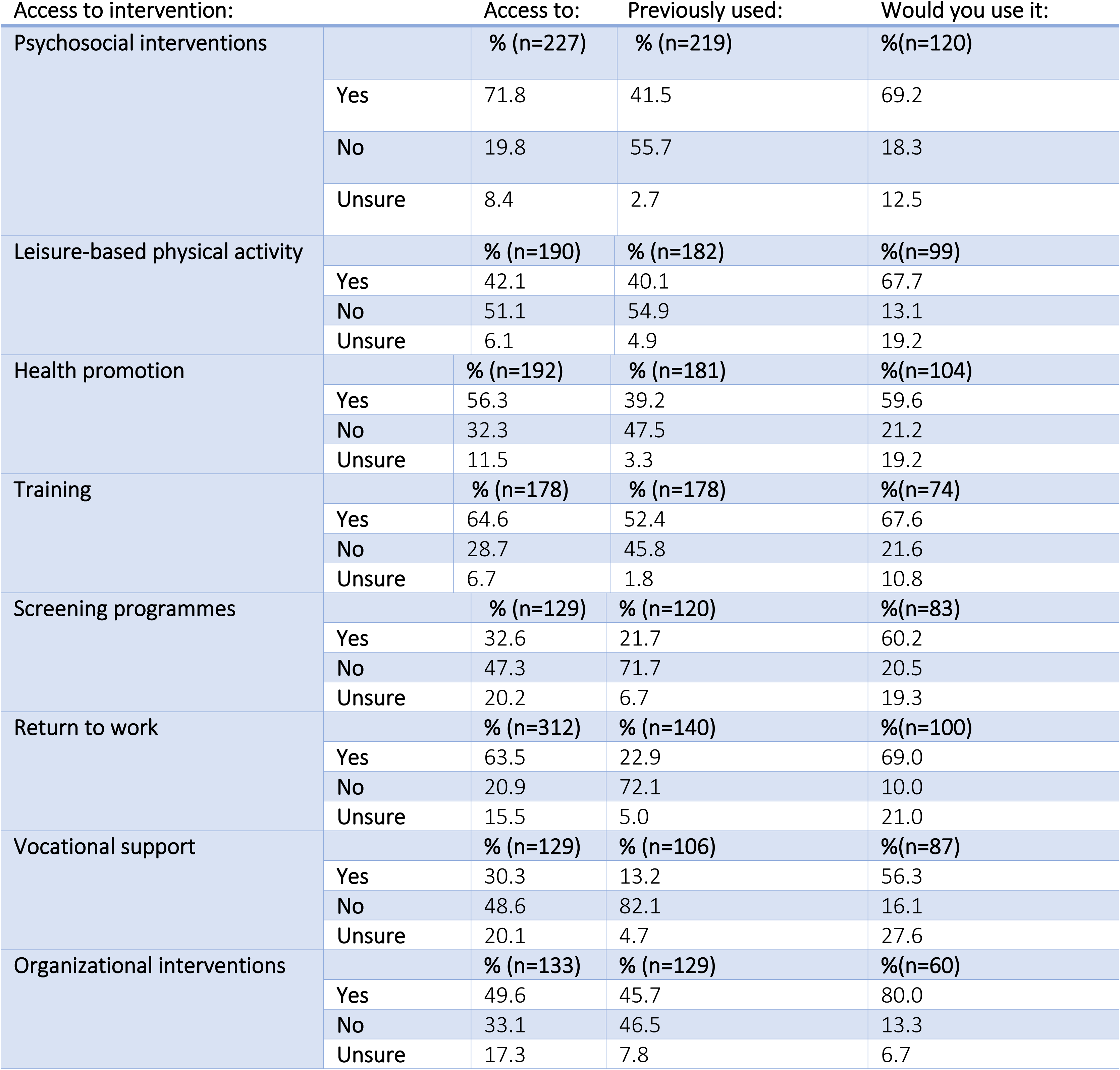
Access to, previous use and willingness to use programs or support at or through work.

We asked respondents who have previously used programs and supports at work to rate the ease of accessing each intervention type. For respondents who had not previously accessed these interventions, we asked them to rate their how easy they perceive access to be. As shown in Table 12, respondents who had previously accessed programs and supports at work reported easier access compared with perceptions about ease of access by those who had not accessed interventions:

**Table 12:** Actual and perceived ease of access.

| Easy to access intervention? |  | Ease of access (if previously used):<br>% (n=91) | Perceived ease of access (if previously not used):<br>% (n=122) |
| --- | --- | --- | --- |
| Psychosocial interventions |  |  |  |
|  | Very easy | 28.5 | 19.7 |
|  | Easy | 47.6 | 36.9 |
|  | Neutral | 13.2 | 20.5 |
|  | Difficult | 8.8 | 8.2 |
|  | Very difficult | 1.1 | 4.9 |
|  | Unsure | 1.1 | 9.8 |
| Leisure-based physical activity |  | % (n=72) | % (n=100) |
|  | Very easy | 40.2 | 20.0 |
|  | Easy | 36.1 | 23.0 |
|  | Neutral | 13.9 | 27.0 |
|  | Difficult | 8.3 | 11.0 |
|  | Very difficult | 0.0 | 4.0 |
|  | Unsure | 1.4 | 15.0 |
| Health promotion |  | % (n=71) | % (n=104) |
|  | Very easy | 32.4 | 8.6 |
|  | Easy | 53.5 | 22.1 |
|  | Neutral | 7.0 | 30.8 |
|  | Difficult | 4.2 | 10.6 |
|  | Very difficult | 2.8 | 8.6 |
|  | Unsure | 0.0 | 19.2 |
| Training |  | % (n=87) | % (n=73) |
|  | Very easy | 35.6 | 9.6 |
|  | Easy | 41.4 | 24.7 |
|  | Neutral | 20.7 | 27.4 |
|  | Difficult | 1.1 | 13.7 |
|  | Very difficult | 1.1 | 4.1 |
|  | Unsure | 0.0 | 20.5 |
| Screening programmes |  | % (n=26) | % (n=86) |
|  | Very easy | 38.5 | 2.3 |
|  | Easy | 34.6 | 19.8 |
|  | Neutral | 7.7 | 26.7 |
|  | Difficult | 11.5 | 19.9 |
|  | Very difficult | 3.8 | 17.4 |
|  | Unsure | 3.8 | 19.8 |
| Return to work |  | % (n=32) | % (n=100) |
|  | Very easy | 15.0 | 16.0 |
|  | Easy | 21.9 | 23.0 |
|  | Neutral | 9.4 | 21.0 |
|  | Difficult | 12.5 | 14.0 |
|  | Very difficult | 6.2 | 7.0 |
|  | Unsure | 0.0 | 19.0 |
| Vocational support |  | % (n=14) | % (n=86) |
|  | Very easy | 21.4 | 2.3 |
|  | Easy | 28.6 | 20.9 |
|  | Neutral | 21.4 | 18.6 |
|  | Difficult | 14.3 | 15.1 |
|  | Very difficult | 7.1 | 10.5 |
|  | Unsure | 7.1 | 32.6 |
| Organizational interventions |  | % (n=59) | % (n=70) |
|  | Extremely easy | 16.9 | 5.7 |
|  | Somewhat easy | 35.6 | 20.0 |
|  | Neither easy nor difficult | 16.9 | 18.6 |
|  | Somewhat difficult | 25.4 | 21.4 |
|  | Extremely difficult | 4.1 | 17.1 |
|  | Unsure | NA | 17.1 |

When asked to rate the level of importance of specific intervention types in supporting mental health at work, the majority of respondents selected extremely or very important across all categories, as described in Table 13.

**Table 13:** To what extent do respondents value the potential interventions for mental health and work?

| *Interventions (N=275)* | **Extremely important %** | **Very important %** | **Moderately important %** | **Slightly or not at all important %** |
| --- | --- | --- | --- | --- |
| **Screening programmes** | 42.2 | 35.4 | 15.6 | 6.8 |
| **Vocational support** | 40.8 | 34.4 | 18.7 | 6.1 |
| **Return to work** | 60.1 | 33.1 | 3.8 | 3.0 |
| **Support for workers with mental health problems (e.g. work accommodations)** | 58.5 | 35.1 | 3.0 | 3.4 |
| **Organizational interventions** | 61.4 | 30.7 | 4.9 | 3.0 |
| **Manager training for mental health** | 62.4 | 29.7 | 4.5 | 3.4 |
| **Access to mental health promotion and prevention (e.g. individual psychosocial interventions, leisure-based physical activity)** | 52.0 | 36.3 | 8.1 | 3.7 |

#### Benefits and Concerns

We assessed perceived benefits and concerns by intervention type as shown in Tables 14 and 15. In terms of benefits (Table 14), being helpful for the worker’s mental health was most highly rated across all intervention types, followed by convenience, affordability for the worker, and being beneficial to the whole organization. For the training and education intervention, the potential to reduce stigma was also rated highly by 19.7% of respondents.

**Table 14:** Perceived benefits of interventions.

| Concerns: | Psychosocial interventions<br>(Total respondents n=222;<br>Total responses n=417)<br>% | Leisure-based physical activity %<br>(Total respondents n=185;<br>Total responses n=240) | Health promotion<br>%<br>(Total respondents n=189;<br>Total responses n=265) | Training<br>%<br>(Total respondents n=172;<br>Total responses n=237) | Screening programmes<br>%<br>(Total respondents n=124;<br>Total responses n=239) | Return to work %<br>(Total respondents n=143;<br>Total responses n=253) | Vocational support<br>%(Total respondents n=106;<br>Total responses n=185) | Organizational interventions<br>%(Total respondents n=130;<br>Total responses n=197) |
| --- | --- | --- | --- | --- | --- | --- | --- | --- |
| Unsure | 0.7 | 9.2 | 7.9 | 4.6 | 3.4 | 4.0 | 10.3 | 5.1 |
| No concerns | 9.4 | 34.2 | 29.4 | 30.4 | 7.5 | 11.5 | 10.3 | 15.7 |
| Other<sup>a</sup> | 7.0 | 9.2 | 7.9 | 7.6 | 4.6 | 5.1 | 2.7 | 8.6 |
| Expensive for employer | 5.3 | 2.9 | 4.9 | 6.8 | 3.8 | 2.8 | 3.8 | 15.7 |
| Expensive for worker | 7.0 | 7.1 | 8.3 | 7.2 | 4.2 | 3.2 | 5.4 | 3.5 |
| Difficult to access | 8.4 | 10.0 | 8.7 | 10.1 | 8.4 | 10.7 | 15.7 | 10.7 |
| Inconvenient | 6.7 | 11.3 | 7.6 | 9.7 | 5.9 | 5.5 | 4.9 | 8.1 |
| Fear of judgment/ stigma | 31.7 | 10.0 | 11.3 | 13.5 | 34.7 | 31.6 | 27.0 | 18.3 |
| Lack of privacy / confidentiality | 24.0 | 6.3 | 14.0 | 10.1 | 27.6 | 25.7 | 20.0 | 14.2 |

<sup>a</sup>Other e.g.: prevents illness; convenient for employer; demonstrates workplace commitment to health; intervention options can be helpful but addressing risk factors for mental health is also important.

**Table 15:**
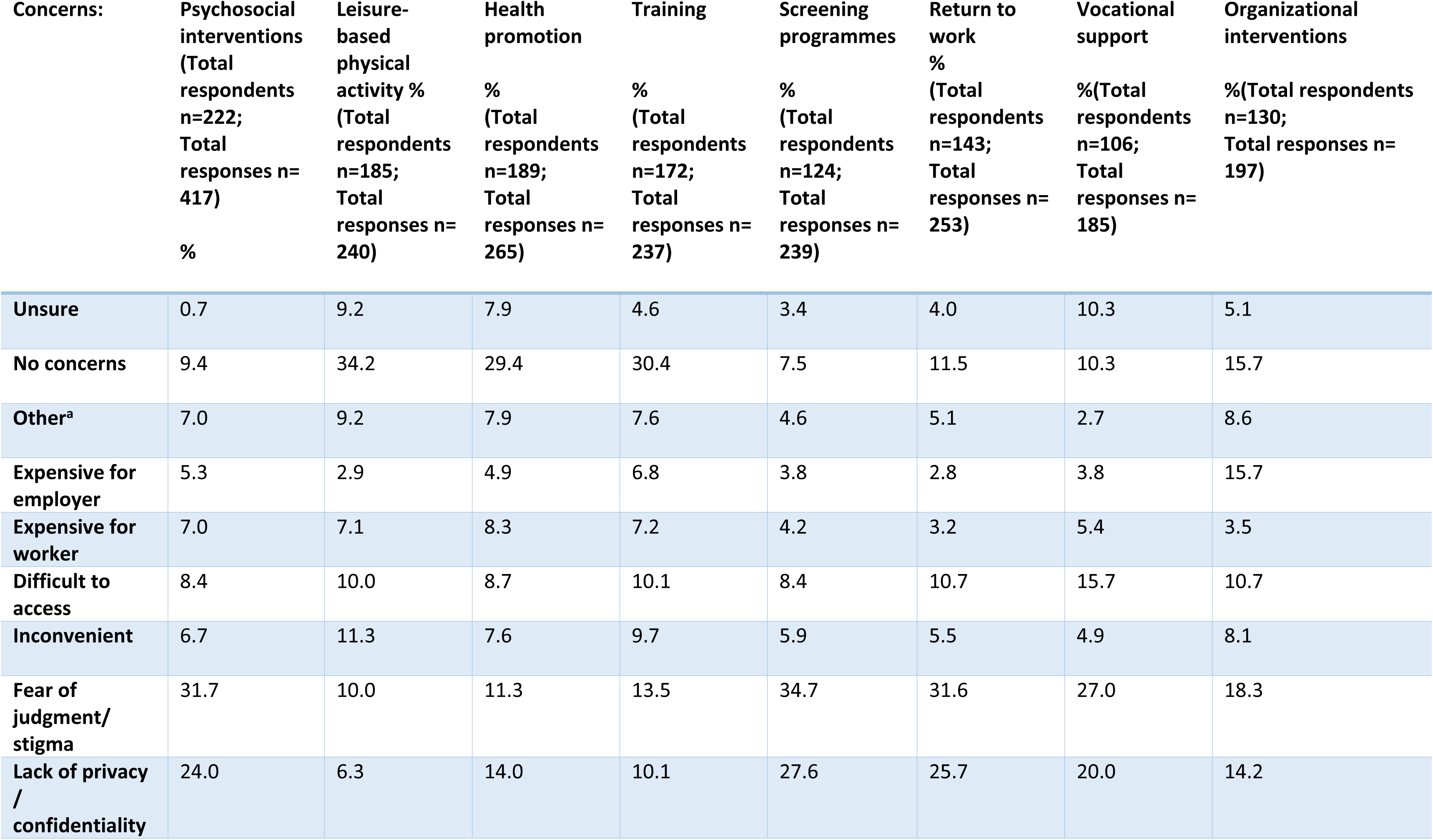
Perceived Concerns of the Interventions.

In qualitative responses describing the benefits of promoting and supporting mental health at work, perceived benefits for workers included improved overall quality of life, well-being, and job satisfaction. It was noted that employees may be empowered and “flourish” through the establishment of a healthy and supportive work environment, as described in the following quotation:

> “Ppl [sic] spend the bulk of their waking hours in work, so I believe that organizations have a duty to go beyond simply keeping places safe and doing no harm, but that by addressing primary concerns at the organizational level employees can thrive. That should be the goal!” (P267, Worker, Woman)

Supporting mental health programs and services at work was also noted as a contributing factor for improving mental health literacy, and, through the provision of information on pathways to care and support, may also improve timely access to care and promote help-seeking among workers. Stigma reduction is also described as a benefit of promoting mental health at work by creating workplace cultures that are open, accepting and are adequately equipped to provide appropriate supports.

Regarding key concerns (Table 15) related to each intervention type, a majority of respondents report no concerns related to physical exercise interventions (34.2%), training and education programs (30.4%) and health promotion programs (29.4%). Across all other intervention types, fear of judgement or stigma, and privacy and confidentiality are rated as the highest concerns.

This reflects concerns raised in the qualitative responses, where respondents were asked to describe any worries they had with regard to promoting and supporting mental health at work, and stigma was raised as a key theme. More specifically, respondents worried that stigma related to mental health would prevent workers from disclosing their mental health challenges and might lead to discrimination or other negative impacts in the workplace.

Ongoing stigma, often associated with low mental health awareness, was also described as a concern. Respondents cautioned that this lack of understanding and awareness may lead to poor treatment of people who are open about their mental health challenges at work, as described below:

> “Because of the stigma around mental health, the people who need help the most are probably the people who are not going to reach out for help. Colleagues and managers should know what to look out for and how to approach someone who they think may be suffering a mental health issue. Seeking help for, and offering assistance to someone who is struggling with a mental health problem, should become as common placed [sic] as seeking help and assisting someone with a physical health problem” (P285, Worker, Woman).

Of additional note, concern related to stigma was highest for screening interventions, with 34.7% of respondents indicating concerns about judgement and stigma and 27.6% reporting concerns with privacy and confidentiality (Table 15).

Respondents were asked to reflect on the potential benefits and harms specifically around mental health screening at work via a short-answer question. In response, participants described several concerns, including privacy, confidentiality and the use of results or data by employers or other parties, including insurance companies. Stigma and discrimination as a result of mental health screening was also raised as a concern with some stating that employees would be reluctant to participate in screening for fear of stigma, ostracization or negative repercussions related to their role at work. Ethical considerations of screening for mental health at work were also raised when pathways to care or services may be limited or unavailable. Some participants questioned the legal implications around duty of care for an organization conducting mental health screening.

Many respondents questioned the appropriateness of the workplace for conducting mental health screening and the validity of an employer having this information about employee mental health, as described below:

> “I don’t believe that the workplace is the setting in which screening should take place. There is a lot of risk for stigma as a result and it is only in very specific circumstances where an employer would need to know the details of this. Workplaces can be a conduit of information, but I would be concerned about them getting ’too close’ to the actual screening.” (P267, Worker, Woman)

Similarly, concerns regarding punitive measures associated with mental health screening at work were also raised by many respondents, as shown in the quotation below:

> “There is evidence that those who share are penalised either by withdrawing promotion, actual demotion or bullied. The anonymity of reporting has to be fiercely guarded.” (P150, Worker, Woman)

Related to these concerns is the suggestion that screening and subsequent interventions or supports should take a population-based approach, avoiding the potential to single out and potentially marginalize individual workers:

> “Screening needs to be done with a view to supporting all workers, and working out who needs what specific additional support. It should not be done as a way of weeding people out, or carrying negative connotations. For example, it would be beneficial to offer mental health awareness and a voluntary stress management training programme. It would be potentially harmful to single out staff members and tell them to complete a stress management programme.” (P285, Manager, Man)

Despite the concerns raised regarding screening for mental health at work, positive themes were identified as well. Possible perceived benefits include the potential for mental health screening at work to lead to prevention and early identification of mental health conditions, thus leading to earlier intervention or care access. Shifts towards a workplace culture that is more supportive and open to discussing issues of mental health and wellbeing was another potential benefit. Proper training and additional supports in the workplace can lead to a workplace culture that actively promotes mental wellbeing were also identified, as described below:

> “Managers should be trained on the sensitivity of mental health information. When an organisation gets it right, like some do, the psychological safety spills into other aspects of work and life of those affected and related.” (P150, Worker, Woman)

#### Organizational Changes

Respondents indicated their awareness of, experiences with, and perceived benefits and concerns about policies or programs aiming to support worker mental health via organizational changes such as those that lead to changes in the working environment, working conditions or work tasks for the purposes of supporting workers’ mental health. We highlight these responses separately as the question types differed slightly from those related to the seven interventions described above.

When asked about perceived benefits of policies and programs to support worker mental health via organizational changes (Table 14) the greatest perceived benefits include being helpful to workers’ mental health (23.9%) and benefitting the whole organization (23.9%).

As extracted from qualitative responses, perceived organizational benefits include improved work quality and productivity among workers, improved workplace culture, greater retention, and reduced absenteeism and presenteeism. Also noted was the anticipated high potential return on investment for businesses or organizations investing in mental health and wellbeing, as described below:

> “A healthy (in a broad sense) workforce is generally a more productive workforce - money invested up-front on preventive strategies (education, stigma reduction, management training), early intervention (HR policies), health care and return to work programs reap the benefits later in numerous ways.” (P275, Manager, Man)

Regarding concerns about introducing policies and programs to support worker mental health via organizational changes (Table 15), the greatest concern among respondents was fear of judgement or stigma (18.3%), followed by cost for the company (15.7%).

Qualitative responses noted a clear desire among respondents for upstream interventions and organizational shifts to support mentally healthy workplaces and to address factors in the workplace that may contribute to poor mental wellbeing. Similarly, respondents raised concerns about organizations creating policies related to workplace mental health without investing in meaningful change to address mental health risk factors. One respondent stated: *“Policies aren’t worth the paper they are printed on without good practice.”* (P273, Manager, Woman). Another cautioned against over pathologizing the workforce by equating challenges with mental health conditions or poor mental wellbeing instead of with the work environment, thus shifting responsibility to the worker instead of the employer:

> “I am concerned that current efforts to support workers mental health focus on building resilience and maximizing worker productivity, rather than putting in systems that ensure better work-life balance.” (P392, Worker, Man)

#### Additional Considerations from Qualitative Responses

We identified other important themes in our analysis of the qualitative responses, as described below.

##### Quality and Appropriateness of Programs and Supports

The quality of mental health programs and supports at work is a concern among respondents, as illustrated in the following quotation that describes reluctance among workers to seek out existing mental health supports:

> “…a lot of employers like higher education institutions rely on commercial and short-term mental health services that use a ’cookie cutter’ approach to CBT and resilience. Staff have noted that these services are not very beneficial in the long run and have learnt not to seek them out.” (P392, Worker, Man)

##### Engagement and Collaboration with Workers

Respondents also emphasized the need for programs and supports that are appropriate and responsive to the needs of workers, calling for collaborative approaches to develop policies and practices that support their mental wellbeing, including by engaging workers with lived experience of mental health conditions, to understand how workplaces can fully support a range of mental health needs.

> “I believe the best mental health initiatives are done in collaboration with workers and their representatives. Managerial approaches to promoting mental health sometimes don’t work so well, but a collaborative approach is best to ensure organizational cultural change.” (P276, Worker, Man)

### Perspectives of Managers

The survey assessed managers’ perspectives and experiences with providing mental health programs and supports at work. All respondents who indicated that they work as a manager were directed to a suite of questions to understand their experience with providing mental health support at work, including via information and referrals and Employee Assistance Programs (EAPs), their perceived self-efficacy regarding providing mental health support to employees, and capacity building needs related to providing mental health support at work.

We asked managers if they have previously provided a referral or information to employees for where or how to access mental health support. 67.3% indicate that they have previously provided a referral or information to employees for where or how to access mental health support. Reasons for not previously having done so include, “It is not related to my job” (46.9%), “It is not available at/through my work” (21.9%), “I do not know what is available at work” (6.3%), or “other” (25%). For ‘other’, reasons provided by managers include not being approached by an employee for support, never having perceived a need among employees for mental health support, and not feeling they know their employees well enough to approach the topic of mental health.

Manager respondents report a high level of confidence to support workers experiencing psychological distress or experiencing mental health conditions, with 38.7% indicating they are very confident and 40.6% stating they are somewhat confident. Despite high levels of confidence selected by managers, when asked to indicate which resources would help to increase their confidence, only 5.0% indicate that they feel sufficiently confident and well-informed^1^. Table 16 describes factors that would help to improve manager confidence to support mental health among employees.

**Table 16:**
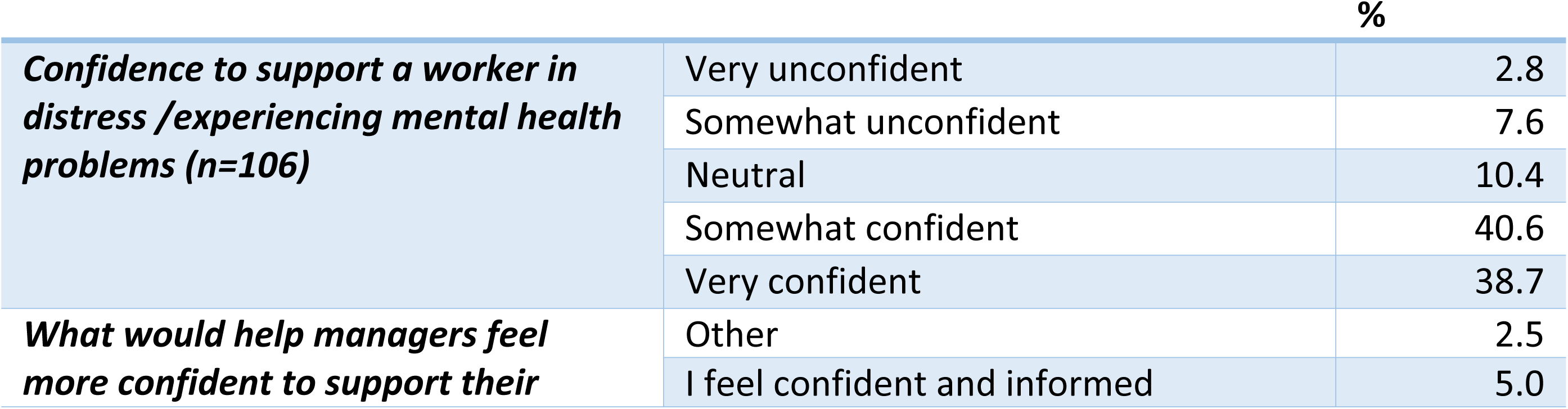
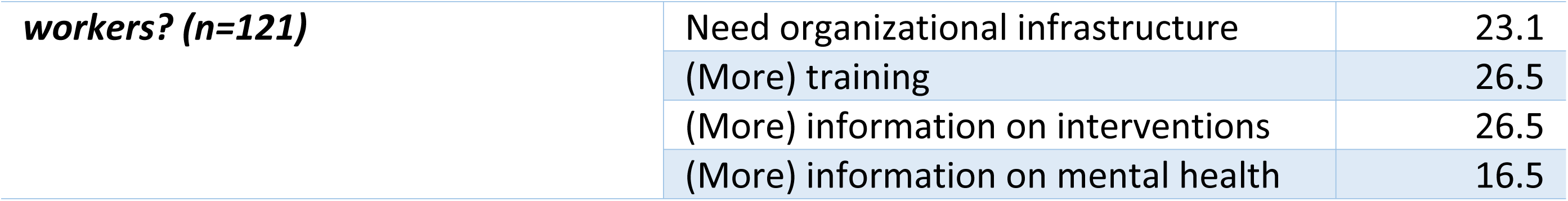
What would help managers to feel more confident to support workers in distress/experiencing mental health issues or challenges.

We also asked managers to identify how easy it would be for them to access training to support knowledge, skills, attitudes, and behaviour to improve mental health of workers when needed. 60.0% indicate that they can access this training when needed, 30.5% have no access to training and 9.5% selected “other”. ‘Other’ responses included some respondents who were unsure about access to training, some managers indicating they can access training but only sporadically, and some stating that training is not yet offered but will be in the future.

Finally, we asked managers to report whether they have experience with EAPs, including through training, developing or managing these programs. A large majority (71.7%) do not have any experience with EAPs, while 28.3% report previous experience.

#### Intervention Specific

Managers have most frequently provided a referral to or information about psychosocial and emotional programs and supports (67.8%) and training and education programs (61.4%). They are least likely to have provided a referral to or information about vocational programs, with 40.0% indicating they had not done so, 33.3% indicating that they had, and 13.0% indicating these programs are not available where they work. Screening was also rated low, with 39.2% indicating they had not, 33.8% indicated they had and 17.6% indicating screening programs are not available where they work. With the exception of screening programs (33.8%) and vocational support programs (26.8%), approximately half of managers indicate that they feel very confident and informed about each intervention type. Managers report the highest level of confidence about psychosocial and emotional supports and programs at 59.2%.

Approximately one quarter of respondents indicate the need for more information about each type of intervention, although the need for more information and training is indicated for vocational support (more information: 39.3%; more training: 23.2%), return to work programs (more information: 30.4%; more training: 15.2%) and screening programs (more information: 32.5%; more training: 20.0%). We also captured manager experiences and perspective with policies and programs to support work mental health through organizational changes. 45.2% of managers have provided a referral or information to employees about accessing or participating in such an intervention, while 28.8% have not and 17.8% indicate that these policies and programs are unavailable in their workplace.

## Discussion

The results of this survey provide a comprehensive understanding of the values and preferences of workers and employers related to the provision of mental health programs and supports at work. Below we present key considerations in reflection of survey findings.

### Organizational Factors

The survey results show that workers place a high level of importance on organizational factors that may either contribute to negative mental health at work or promote mental wellbeing and psychological safety in the workplace. High workload and challenging workflow and organizational culture were most highly rated as having a negative impact on mental health, with qualitative results emphasising the extent of these concerns among survey respondents. This suggests a need for employers and management to improve overall support for workers including via upstream interventions that support factors like work-life balance, healthy interpersonal relationships at work, prevention of extreme overwork, and effective communication between management and employees. These findings are consistent with research that describes the importance of prevention via workplace policies as an essential component of supporting mental health at work (10).

### Training and Capacity Building

Responses by both general workers and managers indicated that more training and capacity building for managers is necessary for providing effective mental health support at work. Although the majority of managers indicate they have some level of confidence to support worker mental health, 95% identified capacity building such as enhanced training and information about mental health issues and interventions as necessary to improve their ability and confidence to provide mental health support to employees.

Almost one half of managers report that they had not provided information or referrals for mental health programs and supports because they don’t perceive it to be part of their job. Others indicate that they have never done so because no employee has ever reached out to them for support. These findings suggest there is a need for enhanced training and capacity building for managers, including by supporting them to take a proactive role in identifying and supporting employee mental health. Evidence supports the importance of management training to promote mental health at work (24). For example, an Australian study found that implementing a mental training program for managers working with fire and rescue workers led to a significant reduction in sick leave by employees and a return on investment of the equivalent of almost £10.00 per pound spent on training (25).

### Stigma

The issue of stigma and related concerns, including experiences of discrimination or punitive measures related to discussing or disclosing mental health concerns, was pervasive throughout the survey results. When describing general values and preferences related to mental health at work, perspectives on specific intervention types, and help-seeking preferences, stigma is at the forefront of concern among workers. Stigma-related issues emerged in several ways. When asked to rate their concerns related to specific intervention types, respondents rated worry about experiencing judgement and stigma and about privacy and confidentiality as top concerns across four intervention types: psychosocial and emotional, screening, return to work, and vocational support programs. Notably, the three interventions for which respondents indicated they had no concerns are interventions that can be delivered at the population level and do not involve singling out specific individuals due to mental health-related concerns: leisure-based physical activity, health promotion, and training and education.

Concerns about stigma are also evident in responses related to help-seeking preferences. Responses regarding preference for sharing mental health concerns suggest a high degree of reticence to openly seek mental health support at work. Respondents prefer to access care from external health services and are most concerned with ensuring that they are able to access programs and supports anonymously and without experiencing negative repercussions at work. This is also reflected in preferences for how to access care-most prefer to access care alone and in-person, and just over one quarter prefer to access support online via digital options. Respondents also report their preference for talking about mental health concerns outside of work or with health services at work, and report high levels of discomfort with speaking with human resource representatives or management.

The substantial influence of stigma and related concerns about experiencing discrimination and other negative repercussions related to being open about or seeking support for mental health at work suggests the potential value of targeted anti-stigma campaigns and interventions to promote a culture of openness and promote help-seeking at work. Evidence suggests that anti-stigma campaigns can improve comfort with disclosing mental health issues, including with employers (26) and can improve supportive behaviour among employees (27). Interventions based on social contact among people without and with lived experience of mental health condition are identified as highly effective in reducing mental health stigma(28). Related to the need for organizational change and supports to promote mental health and wellbeing, enforced policies that protect workers against the negative impacts of stigma related to mental health at work may also be warranted.

### Screening

Respondents show a high level of ambivalence about the use of screening for mental health at work. Quantitative results demonstrate low levels of knowledge, awareness and acceptance of screening programs at work compared with other intervention types. Survey respondents rated screening programs most highly compared with other intervention types for concerns about stigma and judgement, and are screening programs are the least recommended by respondents for implementation at work. The qualitative responses further demonstrate a high level of concern regarding the use of mental health screening at work. Respondents did, however, describe potential positive impacts of screening for mental health at work when done carefully and at a population level, including facilitating early detection and intervention. A US study testing a population-level, opt-in screening intervention with the option to access follow-up care found that the screening program facilitated care uptake among employees (29).

When implemented on a voluntary basis at the population level with adequate privacy and confidentiality measures and available support for those who need it, screening programs may be beneficial in improving early detection and promoting pathways to care.

### Worker Preferences

The survey results provide several key insights into respondents’ preferences regarding specific intervention types. Primarily, in addition to screening programs, respondents report the lowest levels of knowledge, experience and access to return to work and vocational support programs. Manager responses also indicate a low level of experience with and capacity related to these types of interventions. As long-term sickness absence and unemployment are themselves possible consequences of untreated mental health conditions (30), these findings suggest that improved investment in and awareness about such programs and supports among employees and managers would be beneficial.

### Barriers to Access

For those who have not previously accessed mental health programs or supports at work, responses related to perceived ease of access, for which a majority of respondents selected ‘neutral’ or ‘unsure/don’t know’ responses across all intervention types, are low compared with responses from those who have actually accessed these supports. This indicates low levels of knowledge about the availability and accessibility of programs and supports. Previous evidence shows that uptake of workplace mental health programs is often low among workers (31). A Canadian study that implemented mental health awareness training for managers found that improving manager awareness about mental health and mental health interventions increased manager support for workers experiencing mental health conditions and improved employee help-seeking and uptake of mental health supports (32).

Finally, when asked which factors had deterred them from accessing mental health programs and supports at work, respondents indicated that time was the biggest access barrier. Time constraints include workload demands making it difficult for employees to access support, and lack of flexibility in how programs are delivered (e.g. after work hours). Time has previously been identified as a structural barrier to mental health service use by workers experiencing depression (33). Allowing flexibility in work hours to enable workers to participate in mental health programs and supports may thus support increased access to programs and supports.

### Including People with Lived Experience of Mental Health Conditions

The results of the qualitative survey questions include a strong indication of the need for participatory approaches to designing and implementing mental health supports at work, including by involving people with lived or living experience of mental health conditions in the development of organizational policies and programs. The involvement of service users in design and improvement of mental health services is increasingly recognized as essential in the broader mental health field (34). For example, a qualitative study that elicited barriers and facilitators to use of a digital intervention to support workplace mental health (35) points to the importance of engaging with service users in intervention design and implementation planning. This is particularly important when working with multicultural or otherwise diverse teams or when adapting interventions from one setting to another.

## Limitations

The survey sample includes a majority of respondents that are female, reside in Europe and the Americas, are urban-dwelling, highly educated, employed full-time with large businesses or organizations, and have access to benefits (e.g. sick days, vacation time) through their place of work. Over half of respondents work in a role that involves supporting mental health at work, suggesting they are highly familiar with the survey subject matter. Using convenience sampling can bias responses towards people with a pre-existing interest in the topic and with sufficient time and literacy to complete an extensive survey. The survey was available in three languages (English, French and Spanish), which may have limited participation from respondents who do not speak these languages. We believe that this survey offers a comprehensive overview of values and preferences for mental health programs and supports at work among workers internationally, but further research to capture more diverse perspectives could further enrich the findings. Almost 70% of survey respondents have experienced a mental health condition, indicating that this survey has captured the perspectives of people with lived experience of mental health concerns in the workplace. It also reinforces the need for enhanced mental health support at work.

## Conclusions

The results of this study underscore the importance of understanding the perspectives of workers and managers to inform the development and implementation of mental health programs and supports that are acceptable and appropriate for workers themselves, helping to improve uptake and to inform strategies for implementation. The results indicate the need for both population-level and targeted supports that are developed collaboratively with workers, for increased training and capacity building among managers, and for targeted interventions to address the pervasive impact of stigma on perceptions about mental health at work and help-seeking. These results provide a unique and global perspective related to mental health at work from the perspective of workers, employers, and providers of mental health supports at work to inform the development of the WHO Guidelines on Mental Health at Work

## Data Availability

All relevant data are contained with the manuscript and in the Web Annexe to the WHO Guidelines on Mental Health and Work (https://apps.who.int/iris/bitstream/handle/10665/363102/9789240053076-eng.pdf)

## Acknowledgements

The authors would like to sincerely thank everyone who participated in the survey for sharing their time and expertise.

## Footnotes

1 Respondents who indicated they felt ‘very confident’ in the previous question were not directed to this question.

